# BASELINE METABOLIC PROFILING AND RISK OF DEATH FROM COVID-19

**DOI:** 10.1101/2022.01.22.22269691

**Authors:** Glen H. Murata, Allison E. Murata, Heather M. Campbell, Brent Wagner, Benjamin H. Mcmahon, Jenny T. Mao

## Abstract

**Objective:** To derive a predicted probability of death (PDeathLabs) based upon complete value sets for 11 clinical measurements (CM) obtained on patients prior to their diagnosis of coronavirus disease (COVID-19). PDeathLabs is intended for use as a summary metric for baseline metabolic status in multivariate models for COVID-19 death.

**Methods:** Cases were identified through the COVID-19 Shared Data Resource (CSDR) of the Department of Veterans Affairs. The diagnosis required at least one positive nucleic acid amplification test (NAAT). The primary outcome was death within 60 days of the first positive test. We retrieved all values for systolic blood pressure (SBP), diastolic blood pressure (DBP), oxygen saturation (O2SAT), body mass index (BMI), estimated glomerular filtration rate (EGFR), alanine aminotransferase (ALT), serum albumin (ALB), hematocrit (HCT), LDL cholesterol (LDL) hemoglobin A1c (A1C), and HDL cholesterol (HDL) if they were done at least 14 days prior to the NAAT. Clinicians evaluate several attributes of CM that are of critical importance: metabolic control, disease burden, chronicity, refractoriness, tendency to relapse, temporal trends, and lability. We derived 1-3 parameters for each of these attributes: the most recent value (metabolic control); time-weighted average and abnormal area under a severity versus time curve (disease burden); time and number of readings above or below goal (chronicity); longest abnormal cluster and time/number of consecutive readings above goal if the last value was abnormal (refractoriness); number of abnormal clusters (tendency to relapse); long- and short-term changes (temporal trends); and coefficient of variation and mean deviation between consecutive readings (lability). We created computer programs to derive cumulative values for these 13 parameters for all 11 CM as each new value is added. A fitted logistic model was developed for each CM to determine which of the 13 parameters contributed to the risk of death. A main logistic model was developed to determine which of the 13 × 11 = 143 metabolic parameters were independently predictive of death. The resulting model was used to derive PDeathLabs for each patient and the area under its receiver operating characteristic (ROC) curve calculated. Single variable logistic models were also derived for age at diagnosis, the Charlson 2-year (Charl2Yr) and lifetime (CharlEver) scores, and the Elixhauser 2-year (Elix2Yrs) and lifetime (ElixEver) scores. Stata was used to compare the ROCs for PDeathDx and each of the other metrics.

**Results:** On September 30, 2021, there were 347,220 COVID-19 patients in the CSDR. 329,491 (94.9%) patients had CM performed at least 14 days prior to the COVID-19 diagnosis and form the basis for this report. 17,934 (5.44%) died within 60 days of the diagnosis. On the subset regressions, the number of significant parameters ranged from all 13 for SBP to 7 for HDL. 239,393 patients had complete sets of data for developing the main model. Of 143 candidate predictors, 49 parameters were identified as statistically significant, independent predictors of death. The most influential domains were the most recent value, disease burden, temporal trends, and tendency to relapse. The ROC area for PDeathLabs was 0.785 +/- 0.002. No difference was found in the ROC areas of PDeathLabs and age at diagnosis (0.783 +/- 0.002; P = NS). However, the ROC area for PDeathLabs was significantly greater than that of Charl2Yrs (0.704 +/- 0.002; P < 0.001), CharlEver (0.729 +/- 0.002; P < 0.001), Elix2Yrs (0.675 ± 0.002; P < 0.001), and ElixEver (0.707 +/- 0.002; P < 0.001). A poor prognosis was found for chronic systolic hypertension. On the other hand, a higher BMI was protective once SBP, DBP, HDL, LDL and A1C were considered.

**Conclusions:** Our study confirms that parameters derived for 11 CM are significant determinants of COVID-19 death. The most recent value should not be selected over other parameters for multivariate modeling unless there is a physiologic basis for doing so. PDeathLabs has the same discriminating power as age at diagnosis and outperforms comorbidity indices as a summary metric for pre-existing conditions. If validated by others, this approach provides a robust approach to handling CM in multivariate models.

## INTRODUCTION

Many mathematical models have been developed for predicting outcomes of COVID-19 infection (1-5). They have gained popularity because they are very useful for managing patients and allocating scarce resources. Most models are composed of demographic traits, co-morbidity scores, and pre-existing conditions grouped under headings like “malignancy”. Clinical measurements (CM) have not been prominent features of these models. CM provide valuable information about the metabolic status of patients at baseline and include vital signs and routine laboratory tests. Some models have none (1, 5), while others include only one (usually the most recent) of the hundreds of measurements that might be available for each type (2-4). However, there are convincing arguments to expand the use of CM in these prediction rules:

a. A diagnosis informs us that the patient has a certain disease, while CM tell us the burden imposed by its risk factors or the disease itself. The two are not synonymous. For example, one patient with hypertension might have had severely elevated blood pressures (BPs) for decades, while another might be normotensive because of successful treatment. The first would be considered at high risk while the second is “normal”. On the other hand, a patient may not have been given the diagnosis of hypertension even though her BPs are abnormal for a term pregnancy. Classification errors such as these are very common in clinical practice.
b. CM provide some insight into the mechanism of injury. For example, suppose chronic hypoxia is more closely associated with death than acute hypoxia. This observation suggests that cumulative injury over time (such as with secondary pulmonary hypertension) is more influential than the acute metabolic burden imposed by low oxygen levels at presentation.
c. CM add explanatory variables to a prediction model. These components are critical for identifying targets for interventions to reduce the risk. For example, causality might be inferred by a dose-response effect between the CM and risk of the death. The model becomes “hypothesis-generating” and could lead to other studies confirming the mechanism of injury or clinical trials to reduce the risk. Prediction rules rarely define causality in a way that points to a further course of action.

CM are routinely included in periodic health examinations and available for testing in prediction models. They include systolic (SBP) and diastolic blood pressure (DBP), oxygen saturation level (O2Sat), body mass index (BMI), estimated glomerular filtration rate (EGFR), alanine aminotransferase (ALT), serum albumin (ALB), hematocrit (HCT), hemoglobin A1c (A1C), and low-(LDL) and high-density lipoprotein cholesterol (HDL). The major problem is the lack of a systematic approach to selecting the appropriate parameter for each type. There is no evidence that the most recent value of any type is the most influential measurement even though it is common practice to include them in models as a representative value. For example, if microvascular disease mediates the risk posed by diabetes, the most recent A1c is the least suitable measure of tissue glycation. The purpose of this manuscript is to validate a method for summarizing the effects of hundreds of measurements done on each patient for each CM and to evaluate the effects of these parameters on prognosis.

Our approach emulates the way that clinicians interpret CM to reach conclusions about the patient’s metabolic status. Unlike prediction rules, they rarely interpret individual values for CM out of context. In fact, conclusions are often based upon a review of neighboring values or even other domains of the medical record. Moreover, clinicians often render judgements about several attributes when assessing a CM. These attributes include metabolic control, chronicity, disease burden, temporal changes, refractoriness, tendency to relapse, and lability. Each concept is independent of the others and has clinical implications of its own. For example, a diabetic patient may have an A1C that meets a metabolic target, but the entire value set could represent a substantial glycemic burden, indicate a tendency to relapse, and demonstrate a steep, upward trend. In our study, a *parameter* is a variable synthesized from the complete value set for a CM that represents one attribute for that CM. To reach conclusions about metabolic status requires extensive processing of all data for a CM, deriving parameters for each attribute, and testing the association between derived parameters and COVID-19 death. Our computer programs not only derive these parameters but also generate a running summary whenever a new value is added to the set. Thus, the patient’s status relative to any CM can be assessed in detail at any point in time.

## METHODS

Cases were identified through VA’s COVID-19 Shared Data Resource (CSDR). Membership in this registry requires at least one positive nucleic acid amplification test. The primary outcome was death within 60 days of the first positive result. The outcome was retrieved from the CSDR, which assigns a 1 to those who died and 0 otherwise. Data domains in the Corporate Data Warehouse (CDW) were interrogated for all measurements entered ≥ 14 days before the diagnosis of COVID-19. This precaution excludes readings that may have been taken during the pre-symptomatic phases of illness. We retrieved SBP, DBP, O2Sat, heights and weights from the file Vital.VitalSign. The latter were used to calculate BMI. We also retrieved EGFR, ALB, HCT, ALT, A1C, HDL and LDL from the file Chem.PatientLabChem. The following parameters were derived for each of the 11 CMs (above):

a. ***Metabolic control*** requires that the measurement be compared to an external standard; that both be standardized to a common scale; that the value is timely, that it has not already been treated, that enough time has elapsed since the last treatment change to reach a plateau value; and that it is not rapidly increasing or decreasing. Metabolic control is reflected in the most recent value (*Value1*). Our programs provide the option of raw or standardized values and require the user to specify a treatment target. They can be modified to exclude remote values but, for now, include all *Value1s* in the medical record regardless of their timeliness. No attempt was made to assess prior treatments. Temporal changes are handled separately (below).
b. ***Chronicity*** is defined as the total number of days or measurements above target. Each reading was paired with the preceding one. Days between successive abnormal readings were considered above goal; days between normal readings as at goal; days between a normal and abnormal reading as worsening; and days between an abnormal and normal reading as improving. Days above goal were summarized across all successive pairs in the patient’s record (*FUDaysAboveGoal*). *NumAboveGoal* is the total number of measurements exceeding the target value in the patient’s record.
c. ***Disease burden*** is the degree to which an abnormality has caused end-organ damage. For chronic diseases, it is almost always a function of severity and exposure time and expressed as the area under the severity x time curve (AUC). This concept applies to SBP, DBP, BMI, A1c, LDL and HDL. Our software uses a simple trapezoidal estimation technique without interpolation, extrapolation, or curve-fitting and includes all readings from the first to last value. AUC can be large just because the patient is elderly. To eliminate this possibility, our software derives AUC only above the target specified by the user (*AbnAUC)*. Time-weighted averaging eliminates the sampling bias that arises from a tendency to measure a CM more frequently when it is abnormal. The raw average is an over-estimate of severity when abnormal readings are more closely spaced than normal ones. *TimeWtAvg* is defined as: AUC/(Total days of follow-up)
d. ***Refractoriness*** is the repeated failure to achieve goal and is represented by consecutive abnormal values (a “cluster”). Each cluster begins with a change from normal to abnormal value and terminates with the next normal value. The assumption is that an abnormal reading should have triggered an intervention that normalized the measurement. Refractoriness is worse when there are many readings in the cluster, or its duration is long. It can result from the failure to treat, failure of the patient to adhere, failure of the disease to respond, or intolerable side-effects. Our program defines the number of clusters for each patient (*NumCluster*) and a start date, consecutive days above goal (*TimeAboveGoal)*, and consecutive readings above goal (*CtAboveGoal*) for each cluster. The least refractory episode has a *CtAboveGoal* of one, and refractoriness increases as CtAboveGoal increases. *TimeAboveGoal* and *CtAboveGoal* are relevant only for patients with an abnormal CM at the time of COVID diagnosis. The term *MaxClustDays* refers to the longest cluster in the patient’s medical record.
e. ***Relapses*** are the occurrence of abnormal values after being at goal. They can occur once or many times. Relapses are often caused by a steady progression in disease severity (like diabetes) which requires multiple titrations of treatment. The number of relapses is represented by *NumCluster*. The first abnormal cluster is given the value of one – i.e., the person has deteriorated from a previously normal state. Patients with a tendency to relapse have larger values for *NumCluster*. Relapse and refractoriness have different clinical implications. For example, a person can relapse many times, respond quickly to each intervention, and spend little time above goal. On the other hand, a person with refractoriness can have just one cluster that lasts for years.
f. ***Lability*** refers to variations in a CM from one reading to the next. Spontaneous variation can occur when the test is imprecise or when there are fluctuations in the CM itself. Test imprecision may be due to changes in conditions of testing (often behavioral) or variability in the assay. Results of these tests should be confirmed by repeated sampling. On the other hand, physiologic variability results from a complex interplay among multiple control mechanisms and is important in several conditions. For example, labile hypertension is a well-known clinical problem. It has also been shown that hypoglycemia in diabetes results from spontaneous variation in glucose levels and can only rarely be explained by patient behaviors. The coefficient of variation (*CoeffVar*) for each CM is derived from the grand mean (GrandMean) and standard deviation (GrandStD) for all values in the patient record and given by the expression GrandStD/GrandMean. *MeanValDiff* refers to the average of the absolute difference between all consecutive pairs.
g. ***Temporal trends*** refer to changes over time from one value to another and may be long- or short-term. *NetChange* is a measure of long-term trend and refers to the difference in the most recent and first value for each CM. The software generates the mean of the 3 lagging values for each measurement (Lag3Mean). *Lag3Dev* refers to the fractional deviation of the current value from Lag3Mean and is given by the expression: (Value1 – Lag3Mean)/Lag3Mean.

### Statistical methods

In summary, we analyzed the following 13 parameters for each CM: *Value1, FUDaysAboveGoal, NumAboveGoal, AbnAUC, TimeWtAvg, NumClust, TimeAboveGoal, CtAboveGoal, MaxClustDays, CoeffVar, MeanValDiff, NetChange*, and *Lag3Dev*. These values were extracted for each patient from the running summary associated with the most recent value. The default was to use all available readings in the medical record so that metabolic status was defined over the patient’s lifetime.

Because there are 4 vital signs and 7 laboratory tests of interest, the list of covariates consists of 13 × 11 = 143 items. Group differences in categorical variables were tested by chi-square analysis. Group differences in continuous variables were analyzed by student’s t-test and Mann-Whitney U-test. Because there were so many comparisons, univariate analysis is not included in this report but is available upon request.

For each CM, a subset logistic regression was done to identify which of the 13 parameters was predictive of death. A model was fitted to all patients with complete data. A variable was considered significant if the adjusted P-value associated with the coefficient was < 0.05. The subset model was then used to assign a predicted probability of death to each patient. Receiver operating characteristic curve (ROC) analysis was done to determine if that predicted probability discriminated between those who live and died.

All 143 variables were used to generate a main predictive model. Set-wise logistic regression was applied to all patients with a complete data set. The dependent variable was death within 60 days of diagnosis. Modeling started with the set of most recent values. Sets related to different criteria were added in succession, and previous variables were removed if they became insignificant. A variable was considered significant if the adjusted P-value associated with its coefficient was < 0.05. The final model was used to assign a predicted probability of death (PDeathLabs) to each patient based upon all CM. Again, ROC analysis was used to determine if PDeathLabs was able to identify those who died.

## RESULTS

On September 30, 2021, there were 347,220 COVID-19 patients in VA’s COVID-19 Shared Data Resource. Of these, 329,491 patients (94.9%) had CM performed at least 14 days prior to the COVID-19 diagnosis and form the basis for this report. The mean age at the time of diagnosis was 59.1 ± 16.6 years; 85.5% were male; 23.4% were members of a racial minority; 9.2% were Hispanic; 96.4% were veterans; 0.7% were on supplemental oxygen; and 12.2% were current smokers. 9.3% had been fully vaccinated at least 14 days prior to the COVID-19 diagnosis. 21.6% acquired their infections after July 1, 2021 and were presumed to have the delta variant. Overall, 17,934 patients (5.44%) died within 60 days of their diagnosis.

Subset regressions were done for each CM to identify which of the 13 parameters were determinants of the outcome (Appendix). The model did not converge for ALT. As expected, single CM did not distinguish between surviving and dying patients very well, although some ROCs were close to those of co-morbidity scores. Nevertheless, the number of significant parameters ranged from all 13 for SBP to 7 for HDL. *Value1* was significant for 7 CM, *FUDaysAboveGoal* for 9, *NumAboveGoal* for 9, *AbnAUC* for 9, *TimeWtAvg* for 9, *NumClust* for 10, *TimeAboveGoal* for 6, *CtAboveGoal* for 7, *MaxClustDays* for 5, *CoeffVar* for 10, *MeanValDiff* for 10, *NetChange* for 9, and *Lag3Dev* for 7.

239,393 patients (or 70.5% of the cohort) had complete sets of data for developing the main model. Table 1 shows the components of this model. Of 143 candidate predictors, 49 parameters were identified as statistically significant and independent predictors of death. The most influential domains were the most recent value, disease burden, temporal trends, and tendency to relapse. The main model was used to calculate a predicted probability of death (PDeathLabs) for each subject. The ROC area for PDeathLabs was 0.785 ± 0.002. For comparison, single factor logistic models were developed for age at diagnosis, the 2-year Charleson Co-morbidity Index score (Charl2Yrs), lifetime Charleson score (CharlEver), the Elixhauser 2-year score (Elix2Yrs), and lifetime Elixhauser score (ElixEver). Their predicted probabilities were used to derive an ROC area for each. No difference was found in the ROC areas of PDeathLabs and age at diagnosis (0.783 ± 0.002; P > 0.05). However, the ROC area for PDeathLabs was significantly greater than that of Charl2Yrs (0.704 ± 0.002; P < 0.001), CharlEver (0.729 ± 0.002; P < 0.001), Elix2Yrs (0.675 ± 0.002; P < 0.001), and ElixEver (0.707 ± 0.002; P < 0.001). Thus, baseline metabolic measurements outperform co-morbidity scores for pre-existing conditions in predicting COVID-19 deaths.

**TABLE 1:** MULTIVARIATE MODEL FOR COVID-19 DEATH BASED ON BASELINE METABOLIC PROFILES.

| Parameter | Coefficient | Std Error | Z_Score | P_Value | Lower CI | Higher CI |
| --- | --- | --- | --- | --- | --- | --- |
| O2Value1 | -0.0731 | 0.0071 | 10.3300 | 0.0000 | -0.0869 | -0.0592 |
| SBPValue1 | -0.0040 | 0.0009 | -4.5700 | 0.0000 | -0.0058 | -0.0023 |
| DBPValue1 | -0.0082 | 0.0018 | -4.6700 | 0.0000 | -0.0117 | -0.0048 |
| BMIValue1 | -0.0254 | 0.0017 | -15.1600 | 0.0000 | -0.0287 | -0.0221 |
| ALTValue1 | -0.0036 | 0.0010 | -3.4400 | 0.0010 | -0.0056 | -0.0015 |
| AlbValue1 | -0.5831 | 0.0289 | -20.2000 | 0.0000 | -0.6396 | -0.5265 |
| HctValue1 | -0.0115 | 0.0027 | -4.2200 | 0.0000 | -0.0169 | -0.0062 |
| A1cValue1 | 0.0324 | 0.0067 | 4.8200 | 0.0000 | 0.0192 | 0.0456 |
| HDLValue1 | -0.0031 | 0.0008 | -3.8300 | 0.0000 | -0.0047 | -0.0015 |
| O2TimeWtAvg | -0.1485 | 0.0092 | -16.2000 | 0.0000 | -0.1664 | -0.1305 |
| SBPTimeWtAvg | 0.0297 | 0.0013 | 22.2300 | 0.0000 | 0.0271 | 0.0323 |
| DBPTimeWtAvg | -0.0424 | 0.0021 | -19.9300 | 0.0000 | -0.0466 | -0.0383 |
| EGFRTIMEWtAvg | -0.0131 | 0.0009 | -15.3100 | 0.0000 | -0.0148 | -0.0115 |
| ALTTimeWtAvg | -0.0186 | 0.0021 | -8.8000 | 0.0000 | -0.0228 | -0.0145 |
| LDLTimeWtAvg | -0.0074 | 0.0006 | -11.7900 | 0.0000 | -0.0086 | -0.0062 |
| O2AbnAUC | -0.0001 | 0.0000 | -7.6200 | 0.0000 | -0.0001 | -0.0001 |
| EGFRAbnAUC | 0.0000 | 0.0000 | 3.9700 | 0.0000 | 0.0000 | 0.0000 |
| LDLAbnAUC | 0.0000 | 0.0000 | 5.3500 | 0.0000 | 0.0000 | 0.0000 |
| ALTAbnAUC | 0.0000 | 0.0000 | 7.5600 | 0.0000 | 0.0000 | 0.0000 |
| SBPNumAboveGoal | -0.0010 | 0.0002 | -5.5200 | 0.0000 | -0.0014 | -0.0006 |
| BMINumAboveGoal | -0.0018 | 0.0004 | -4.9200 | 0.0000 | -0.0025 | -0.0011 |
| LDLNumAboveGoal | 0.0075 | 0.0021 | 3.5500 | 0.0000 | 0.0033 | 0.0116 |
| HDLFUDaysBelowGoal | 0.0000 | 0.0000 | 3.5300 | 0.0000 | 0.0000 | 0.0000 |
| AlbCtBelowGoal | -0.0128 | 0.0043 | -2.9400 | 0.0030 | -0.0213 | -0.0043 |
| LDLTimeAboveGoal | 0.0000 | 0.0000 | -1.9900 | 0.0460 | 0.0000 | 0.0000 |
| HDLTimeBelowGoal | 0.0000 | 0.0000 | -3.3100 | 0.0010 | 0.0000 | 0.0000 |
| EGFRMeanValDiff | 0.0126 | 0.0026 | 4.8700 | 0.0000 | 0.0075 | 0.0176 |
| AlbMeanValDiff | -0.4072 | 0.1128 | -3.6100 | 0.0000 | -0.6282 | -0.1862 |
| SBPCoeffVar | 0.0643 | 0.0036 | 17.6700 | 0.0000 | 0.0572 | 0.0715 |
| BMICoeffVar | 0.0152 | 0.0024 | 6.2600 | 0.0000 | 0.0104 | 0.0199 |
| AlbCoeffVar | 0.0136 | 0.0030 | 4.5500 | 0.0000 | 0.0077 | 0.0194 |
| HctCoeffVar | 0.0137 | 0.0023 | 6.0500 | 0.0000 | 0.0092 | 0.0181 |
| SBPNumClust | 0.0021 | 0.0004 | 5.0300 | 0.0000 | 0.0013 | 0.0029 |
| EGFRNumClust | 0.0112 | 0.0033 | 3.3700 | 0.0010 | 0.0047 | 0.0177 |
| ALTNumClust | 0.0254 | 0.0059 | 4.3500 | 0.0000 | 0.0140 | 0.0369 |
| AlbNumClust | -0.0227 | 0.0074 | -3.0800 | 0.0020 | -0.0372 | -0.0082 |
| HctNumClust | -0.0150 | 0.0040 | -3.7800 | 0.0000 | -0.0228 | -0.0072 |
| LDLNumClust | 0.0304 | 0.0064 | 4.7700 | 0.0000 | 0.0179 | 0.0429 |
| A1cNumClust | 0.0423 | 0.0067 | 6.3300 | 0.0000 | 0.0292 | 0.0554 |
| O2Lag3Dev | 3.8382 | 0.4806 | 7.9900 | 0.0000 | 2.8962 | 4.7803 |
| SBPLag3Dev | 0.3222 | 0.1195 | 2.7000 | 0.0070 | 0.0879 | 0.5565 |
| DBPLag3Dev | 0.3167 | 0.1249 | 2.5300 | 0.0110 | 0.0718 | 0.5616 |
| EGFRLag3Dev | -0.1797 | 0.0531 | -3.3800 | 0.0010 | -0.2839 | -0.0756 |
| LDLLag3Dev | 0.1140 | 0.0317 | 3.6000 | 0.0000 | 0.0519 | 0.1761 |
| O2NetChange | 0.0098 | 0.0039 | 2.5100 | 0.0120 | 0.0022 | 0.0175 |
| AlbNetChange | 0.1491 | 0.0234 | 6.3800 | 0.0000 | 0.1032 | 0.1949 |
| HctNetChange | -0.0161 | 0.0025 | -6.4100 | 0.0000 | -0.0210 | -0.0112 |
| LDLNetChange | -0.0006 | 0.0002 | -2.3700 | 0.0180 | -0.0011 | -0.0001 |
| A1cNetChange | 0.0260 | 0.0063 | 4.1000 | 0.0000 | 0.0136 | 0.0384 |
| Constant | 23.4159 | 0.7186 | 32.5800 | 0.0000 | 22.0075 | 24.8244 |

## DISCUSSION

In this manuscript, we propose a novel method for handling pre-existing findings from vital signs and laboratory tests in models predicting COVID-19 death. Our observations confirm that they should be included because they have independent prognostic significance, generate insights into the mechanism of action, and may even be targets for interventions to mitigate the risk. By including explanatory variables, models can become hypothesis-generating and lead to future studies validating the suspected pathogenesis or trials of interventions targeting the abnormality itself.

All 13 parameters were identified as predictors of death in the subset models. These observations provide some justification to the approach that clinicians use to evaluate metabolic status. It is comforting to know that our parameters and the approach used in clinical practice converge upon a common set of criteria. Our main model showed that many parameters besides the most recent value were significant contributors to COVID-19 death. For 2 CM, disease burden contributed to the prediction while the most recent value did not. For 6 others, both current metabolic control and the long-term parameters were independently predictive of death. This finding suggests that CM have chronic and acute effects on the likelihood of recovery from COVID-19 infection and that the former should routinely be considered for prediction models. Our results confirm the importance of CM parameters for COVID-19 prognosis, validate the attributes that they represent, and point out the hazards of using the most recent value as the most representative one.

This study illustrates the complex interactions that become apparent when multiple CM are studied in context. Higher values for O2Value1 and O2TimeWtAvg in our main model independently decreased the risk of death. This observation implies that the metabolic consequences of acute hypoxia and the burden imposed by chronic hypoxia create a substrate for COVID-19 injury. The latter may have done so through secondary pulmonary hypertension or may have simply been a marker for severe lung disease. Of 4 BP parameters, only SBPTimeWtAvg was a risk factor for COVID-19 death. Higher values for DBPValue1 and DBPTimeWtAvg were protective. This paradoxical effect might be explained by the fact that, for a given level of risk posed by SBP, a higher DBP is associated with a lower pulse pressure, reduced systolic wall stress, and better coronary perfusion. In a previous study, we showed that hypertension is the most important independent risk factor for COVID-19 death (6). This analysis suggests that the most important mechanism is long-term elevation of SBP. BMI and obesity are considered risk factors for COVID-related death (2, 5). However, we found that higher values for BMIValue1 in the main model lowered the risk of death. This discrepancy might be explained by the fact that we controlled for the mediators of injury in obesity (SBP, DBP, A1C, HDL, and LDL). Once these factors are considered, a higher BMI may only be a marker of better nutrition. This finding illustrates the hazards of drawing conclusions from incompletely specified models. Serum albumin and hematocrit are indicators of nutritional status or catabolism and are powerful predictors of recovery from acute illness. On the other hand, they do not produce cumulative organ injury over years. Accordingly, ALBValue1 and HCTValue1 were included in the main model but not their long-term parameters. A high value for A1CValue1 was identified as a risk factor while measures of disease burden were not. Thus, recent hyperglycemia is deleterious in COVID-19 infection but microvascular injury developing over decades may not play that much of a role. Likewise, HDLValue1 had a protective effect while the long-term parameters did not. Finally, we found that time-weighted averages and abnormal AUC were both included as measures of disease burden for EGFR, ALT, LDL, and O2SAT. For the last 3, the effects were in opposite directions. These results imply that abnormal AUC provides information about risk beyond the time weighted average. This study shows that there is no justification for selecting one arbitrary parameter of a CM for modeling over the others unless there is a physiologic basis for doing so.

We also found that temporal instability in a CM – whether reflected in long- or short-term trends, coefficient of variation, or tendency to relapse – had significant effects on prognosis. The direction and magnitude of this effect varied from CM to CM and, at times, were unexpected. Future studies should be done to identify the mechanisms by which fluctuations in CM affect recovery from COVID-19 infection. Until then, it is reasonable to include temporal changes of CM in models of COVID-19 death. This recommendation is consistent with the way that clinicians prioritize treatment of abnormal values. For example, many would consider a patient whose A1c rapidly increased from 5.0 to 8.5 to have a poorer prognosis than one whose A1c decreased from 11.0 to 9.0.

As expected, measures of chronicity added little to the model once disease burden was considered. The minimal contribution from refractoriness suggests that clustering adds very little to the prognosis compared to the same abnormalities scattered over the patient’s timeline. Nevertheless, it remains a useful clinical concept because it identifies patients for whom further treatment may be futile.

We calculated a predicted probability of death (PDeathLabs) based upon complete value sets for 11 CM. The ROC area indicated that vital signs and laboratory findings are powerful predictors of outcomes. In fact, the ROC areas for PDeathLabs and age at diagnosis were equivalent, while PDeathLabs was consistently superior to Charl2Yrs, CharlEver, Elix2Yrs, and ElixEver. In summary, this term was a very convenient way to summarize millions of observations for CM done on hundreds of thousands of patients. Its greatest utility would be as a covariate in a parent model containing many other domains.

We did not include age in our model for PDeathLabs for several reasons. First, it was intended to represent pre-existing clinical measurements in an overarching model containing multiple domains. One domain was comprised of demographic characteristics including age at diagnosis. Next, we did not want age to displace clinical parameters in the model highly correlated with age such as blood pressure or weight. To understand the mechanisms that lead to a fatal outcome, explanatory variables should take precedence over disease markers. They may even be targets for an intervention. Finally, models based upon demographics do not support decision-making at the point of care. For example, a physician might feel comfortable withdrawing care if the patient had refractory hypoxemia or terminal renal failure – but not simply because the patient was old. Clinicians make recommendations based upon an assessment of the underlying conditions. It is the patient’s prerogative to decide what is appropriate based upon age and quality of life.

Of course, our conclusions are limited to patients with characteristics like the veteran population. Moreover, this study was confined to patients with chronic illnesses that required long-term follow-up and periodic health screening. It is unclear if our conclusions are applicable to patients without such indications. Further studies should be done on other populations and disease states before the method should be widely applied. If validated by others, our method could revolutionize the way in which clinic measurements are handled in multivariate models.

## Data Availability

All data produced in the present study are available upon reasonable request to the authors.

## APPENDIX A: MULTIVARIATE MODEL FOR COVID-19 DEATH USING O2 PARAMETERS

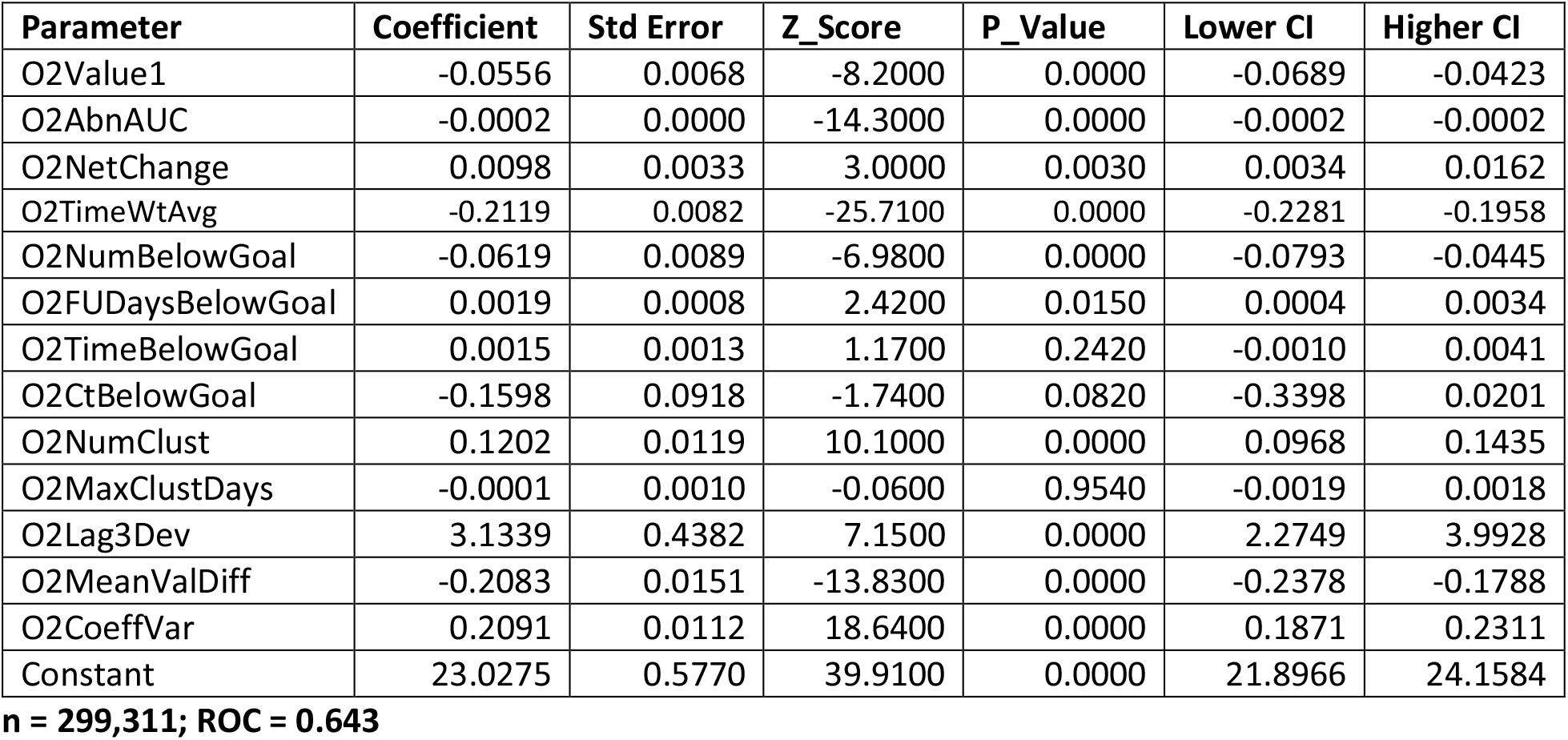

| Parameter | Coefficient | Std Error | Z_Score | P_Value | Lower CI | Higher CI |
| --- | --- | --- | --- | --- | --- | --- |
| O2Value1 | -0.0556 | 0.0068 | -8.2000 | 0.0000 | -0.0689 | -0.0423 |
| O2AbnAUC | -0.0002 | 0.0000 | -14.3000 | 0.0000 | -0.0002 | -0.0002 |
| O2NetChange | 0.0098 | 0.0033 | 3.0000 | 0.0030 | 0.0034 | 0.0162 |
| O2TimeWtAvg | -0.2119 | 0.0082 | -25.7100 | 0.0000 | -0.2281 | -0.1958 |
| O2NumBelowGoal | -0.0619 | 0.0089 | -6.9800 | 0.0000 | -0.0793 | -0.0445 |
| O2FUDaysBelowGoal | 0.0019 | 0.0008 | 2.4200 | 0.0150 | 0.0004 | 0.0034 |
| O2TimeBelowGoal | 0.0015 | 0.0013 | 1.1700 | 0.2420 | -0.0010 | 0.0041 |
| O2CtBelowGoal | -0.1598 | 0.0918 | -1.7400 | 0.0820 | -0.3398 | 0.0201 |
| O2NumClust | 0.1202 | 0.0119 | 10.1000 | 0.0000 | 0.0968 | 0.1435 |
| O2MaxClustDays | -0.0001 | 0.0010 | -0.0600 | 0.9540 | -0.0019 | 0.0018 |
| O2Lag3Dev | 3.1339 | 0.4382 | 7.1500 | 0.0000 | 2.2749 | 3.9928 |
| O2MeanValDiff | -0.2083 | 0.0151 | -13.8300 | 0.0000 | -0.2378 | -0.1788 |
| O2CoeffVar | 0.2091 | 0.0112 | 18.6400 | 0.0000 | 0.1871 | 0.2311 |
| Constant | 23.0275 | 0.5770 | 39.9100 | 0.0000 | 21.8966 | 24.1584 |

## APPENDIX B: MULTIVARIATE MODEL FOR COVID-19 DEATH USING SBP PARAMETERS

**n = 299,311; ROC = 0.643** **APPENDIX B: MULTIVARIATE MODEL FOR COVID-19 DEATH USING SBP PARAMETERS**
| Parameter | Coefficient | Std Error | Z_Score | P_Value | Lower CI | Higher CI |
| --- | --- | --- | --- | --- | --- | --- |
| SBPValue1 | -0.0039 | 0.0009 | -4.5500 | 0.0000 | -0.0056 | -0.0022 |
| SBPAbnAUC | 0.0000 | 0.0000 | -7.1200 | 0.0000 | 0.0000 | 0.0000 |
| SBPNetChange | -0.0060 | 0.0004 | -13.9000 | 0.0000 | -0.0069 | -0.0052 |
| SBPTimeWtAvg | 0.0092 | 0.0013 | 7.0300 | 0.0000 | 0.0066 | 0.0118 |
| SBPNumAboveGoal | -0.0011 | 0.0002 | -7.3200 | 0.0000 | -0.0014 | -0.0008 |
| SBPFUDaysAboveGoal | 0.0004 | 0.0000 | 18.7000 | 0.0000 | 0.0004 | 0.0005 |
| SBPTimeAboveGoal | -0.0002 | 0.0001 | -2.6100 | 0.0090 | -0.0003 | 0.0000 |
| SBPCtAboveGoal | 0.0100 | 0.0035 | 2.8200 | 0.0050 | 0.0030 | 0.0169 |
| SBPNumClust | 0.0039 | 0.0004 | 10.6800 | 0.0000 | 0.0032 | 0.0046 |
| SBPMaxClustDays | -0.0003 | 0.0000 | -12.4200 | 0.0000 | -0.0004 | -0.0003 |
| SBPLag3Dev | 0.6649 | 0.0879 | 7.5600 | 0.0000 | 0.4925 | 0.8372 |
| SBPMeanValDiff | -0.0152 | 0.0028 | -5.5300 | 0.0000 | -0.0207 | -0.0098 |
| SBPCoeffVar | 0.1830 | 0.0035 | 51.5700 | 0.0000 | 0.1760 | 0.1899 |
| Constant | -5.4780 | 0.1413 | -38.7600 | 0.0000 | -5.7550 | -5.2010 |

## APPENDIX C: MULTIVARIATE MODEL FOR COVID-19 DEATH USING DBP PARAMETERS

**n = 310,800; R = 0.711** **APPENDIX C: MULTIVARIATE MODEL FOR COVID-19 DEATH USING DBP PARAMETERS**
| Parameter | Coefficient | Std Error | Z_Score | P_Value | LowCI | HighCI |
| --- | --- | --- | --- | --- | --- | --- |
| DBPValue1 | -0.0243 | 0.0015 | -15.8600 | 0.0000 | -0.0273 | -0.0213 |
| DBPAbnAUC | 0.0000 | 0.0000 | 3.4200 | 0.0010 | 0.0000 | 0.0000 |
| DBPNetChange | -0.0137 | 0.0007 | -19.1300 | 0.0000 | -0.0151 | -0.0123 |
| DBPTimeWtAvg | -0.0329 | 0.0019 | -17.5200 | 0.0000 | -0.0366 | -0.0292 |
| DBPNumAboveGoal | -0.0042 | 0.0007 | -6.4600 | 0.0000 | -0.0055 | -0.0029 |
| DBPFUDaysAboveGoal | 0.0002 | 0.0001 | 3.7800 | 0.0000 | 0.0001 | 0.0004 |
| DBPTimeAboveGoal | -0.0002 | 0.0002 | -0.8800 | 0.3760 | -0.0005 | 0.0002 |
| DBPCtAboveGoal | 0.0501 | 0.0133 | 3.7700 | 0.0000 | 0.0240 | 0.0761 |
| DBPNumClust | 0.0101 | 0.0011 | 9.3300 | 0.0000 | 0.0080 | 0.0122 |
| DBPMaxClustDays | -0.0002 | 0.0001 | -3.3300 | 0.0010 | -0.0004 | -0.0001 |
| DBPLag3Dev | 1.5364 | 0.0878 | 17.4900 | 0.0000 | 1.3642 | 1.7085 |
| DBPMeanValDiff | -0.1277 | 0.0052 | -24.5500 | 0.0000 | -0.1379 | -0.1175 |
| DBPCoeffVar | 0.1646 | 0.0037 | 44.8700 | 0.0000 | 0.1574 | 0.1718 |
| Constant | 0.5617 | 0.1176 | 4.7800 | 0.0000 | 0.3312 | 0.7922 |

## APPENDIX D: MULTIVARIATE MODEL FOR COVID-19 DEATH USING BMI PARAMETERS

**n = 310,792; ROC = 0.705** APPENDIX D: MULTIVARIATE MODEL FOR COVID-19 DEATH USING BMI PARAMETERS
| Parameter | Coefficient | Std Error | Z_Score | P_Value | Lower CI | Higher CI |
| --- | --- | --- | --- | --- | --- | --- |
| BMIValue1 | -0.0547 | 0.0053 | -10.3200 | 0.0000 | -0.0650 | -0.0443 |
| BMIAbnAUC | 0.0000 | 0.0000 | 13.3800 | 0.0000 | 0.0000 | 0.0000 |
| BMINetChange | -0.0338 | 0.0028 | -11.8800 | 0.0000 | -0.0394 | -0.0282 |
| BMITimeWtAvg | -0.0017 | 0.0058 | -0.3000 | 0.7630 | -0.0131 | 0.0096 |
| BMINumAboveGoal | -0.0025 | 0.0006 | -4.0600 | 0.0000 | -0.0037 | -0.0013 |
| BMIFUDaysAboveGoal | 0.0001 | 0.0000 | 6.8900 | 0.0000 | 0.0001 | 0.0002 |
| BMITimeAboveGoal | -0.0001 | 0.0000 | -5.9800 | 0.0000 | -0.0001 | 0.0000 |
| BMICtAboveGoal | 0.0053 | 0.0008 | 7.0300 | 0.0000 | 0.0038 | 0.0068 |
| BMINumClust | 0.0493 | 0.0050 | 9.8100 | 0.0000 | 0.0395 | 0.0592 |
| BMIMaxClustDays | 0.0000 | 0.0000 | -0.8400 | 0.4010 | -0.0001 | 0.0000 |
| BMILag3Dev | -0.1520 | 0.1522 | -1.0000 | 0.3180 | -0.4503 | 0.1462 |
| BMIMeanValDiff | -0.2505 | 0.0165 | -15.1800 | 0.0000 | -0.2828 | -0.2181 |
| BMICoeffVar | 0.0596 | 0.0027 | 21.7500 | 0.0000 | 0.0542 | 0.0650 |
| Constant | -1.4756 | 0.0728 | -20.2600 | 0.0000 | -1.6184 | -1.3329 |

## APPENDIX E: MULTIVARIATE MODEL FOR COVID-19 DEATH USING EGFR PARAMETERS

**n = 296,942; ROC = 0.650** APPENDIX E: MULTIVARIATE MODEL FOR COVID-19 DEATH USING EGFR PARAMETERS
| Parameter | Coefficient | Std Error | Z_Score | P_Value | Lower CI | Higher CI |
| --- | --- | --- | --- | --- | --- | --- |
| EGFRValue1 | -0.0091 | 0.0011 | -8.3600 | 0.0000 | -0.0113 | -0.0070 |
| EGFRAbnAUC | 0.0000 | 0.0000 | 5.5800 | 0.0000 | 0.0000 | 0.0000 |
| EGFRNetChange | 0.0064 | 0.0006 | 10.3000 | 0.0000 | 0.0051 | 0.0076 |
| EGFRTimeWtAvg | -0.0096 | 0.0012 | -8.0400 | 0.0000 | -0.0119 | -0.0072 |
| EGFRNumBelowGoal | -0.0054 | 0.0009 | -5.9700 | 0.0000 | -0.0072 | -0.0036 |
| EGFRFUDaysBelowGoal | 0.0002 | 0.0000 | 5.7200 | 0.0000 | 0.0001 | 0.0002 |
| EGFRTimeBelowGoal | -0.0001 | 0.0000 | -3.2600 | 0.0010 | -0.0001 | 0.0000 |
| EGFRctBelowGoal | 0.0070 | 0.0010 | 6.9500 | 0.0000 | 0.0050 | 0.0090 |
| EGFRNumClust | 0.0879 | 0.0037 | 23.7900 | 0.0000 | 0.0807 | 0.0952 |
| EGFRMaxClustDays | 0.0000 | 0.0000 | -0.3800 | 0.7070 | -0.0001 | 0.0001 |
| EGFRLag3Dev | -0.0054 | 0.0568 | -0.0900 | 0.9250 | -0.1166 | 0.1059 |
| EGFRMeanValDiff | 0.0058 | 0.0020 | 2.9300 | 0.0030 | 0.0019 | 0.0097 |
| EGFRCoeffVar | 0.0198 | 0.0010 | 18.8900 | 0.0000 | 0.0178 | 0.0219 |
| Constant | -2.0081 | 0.0567 | -35.4100 | 0.0000 | -2.1192 | -1.8970 |

## APPENDIX F: MULTIVARIATE MODEL FOR COVID-19 DEATH USING ALB PARAMETERS

**n = 294,314; ROC = 0.691** **APPENDIX F: MULTIVARIATE MODEL FOR COVID-19 DEATH USING ALB PARAMETERS**
| Parameter | Coefficient | Std Error | Z_Score | P_Value | Lower CI | HigherCI |
| --- | --- | --- | --- | --- | --- | --- |
| AlbValue1 | -1.2062 | 0.0428 | -28.1500 | 0.0000 | -1.2902 | -1.1223 |
| AlbAbnAUC | -0.0015 | 0.0002 | -6.7600 | 0.0000 | -0.0019 | -0.0010 |
| AlbNetChange | 0.1897 | 0.0252 | 7.5200 | 0.0000 | 0.1403 | 0.2392 |
| AlbTimeWtAvg | -0.1302 | 0.0458 | -2.8500 | 0.0040 | -0.2199 | -0.0405 |
| AlbNumBelowGoal | -0.0179 | 0.0024 | -7.4900 | 0.0000 | -0.0226 | -0.0132 |
| AlbFUDaysBelowGoal | 0.0007 | 0.0002 | 3.0100 | 0.0030 | 0.0002 | 0.0012 |
| AlbTimeBelowGoal | 0.0002 | 0.0002 | 1.3500 | 0.1760 | -0.0001 | 0.0005 |
| AlbCtBelowGoal | -0.0408 | 0.0060 | -6.8500 | 0.0000 | -0.0525 | -0.0291 |
| AlbNumClust | 0.0737 | 0.0086 | 8.6000 | 0.0000 | 0.0569 | 0.0905 |
| AlbMaxClustDays | -0.0002 | 0.0003 | -0.8900 | 0.3730 | -0.0008 | 0.0003 |
| AlbLag3Dev | 0.8988 | 0.1077 | 8.3400 | 0.0000 | 0.6877 | 1.1100 |
| AlbMeanValDiff | -1.3064 | 0.0959 | -13.6300 | 0.0000 | -1.4943 | -1.1185 |
| AlbCoeffVar | 0.0633 | 0.0029 | 22.1700 | 0.0000 | 0.0577 | 0.0689 |
| Constant | 2.3914 | 0.1173 | 20.3800 | 0.0000 | 2.1615 | 2.6214 |

## APPENDIX G: MULTIVARIATE MODEL FOR COVID-19 DEATH USING HCT VALUES

**n = 286,066; ROC = 0.693** **APPENDIX G: MULTIVARIATE MODEL FOR COVID-19 DEATH USING HCT VALUES**
| Parameter | Coefficient | Std Error | Z_Score | P_Value | Lower CI | Higher CI |
| --- | --- | --- | --- | --- | --- | --- |
| HctValue1 | -0.0312 | 0.0042 | -7.4900 | 0.0000 | -0.0393 | -0.0230 |
| HctAbnAUC | -0.0001 | 0.0000 | -3.8300 | 0.0000 | -0.0001 | 0.0000 |
| HctNetChange | -0.0437 | 0.0029 | -15.1900 | 0.0000 | -0.0493 | -0.0380 |
| HctTimeWtAvg | -0.0249 | 0.0045 | -5.5500 | 0.0000 | -0.0337 | -0.0161 |
| HctNumBelowGoal | -0.0065 | 0.0009 | -6.9500 | 0.0000 | -0.0083 | -0.0046 |
| HctFUDaysBelowGoal | 0.0005 | 0.0001 | 4.4100 | 0.0000 | 0.0003 | 0.0008 |
| HctTimeBelowGoal | 0.0002 | 0.0001 | 2.2000 | 0.0280 | 0.0000 | 0.0005 |
| HctCtBelowGoal | -0.0100 | 0.0025 | -3.9200 | 0.0000 | -0.0149 | -0.0050 |
| HctNumClust | 0.0406 | 0.0049 | 8.2600 | 0.0000 | 0.0310 | 0.0503 |
| HctMaxClustDays | -0.0004 | 0.0002 | -2.8100 | 0.0050 | -0.0007 | -0.0001 |
| HctLag3Dev | 0.4425 | 0.1106 | 4.0000 | 0.0000 | 0.2258 | 0.6592 |
| HctMeanValDiff | -0.0812 | 0.0107 | -7.5600 | 0.0000 | -0.1023 | -0.0602 |
| HctCoeffVar | 0.0764 | 0.0023 | 32.9700 | 0.0000 | 0.0718 | 0.0809 |
| Constant | -0.9129 | 0.1121 | -8.1400 | 0.0000 | -1.1327 | -0.6931 |

## APPENDIX H: MULTIVARIATE MODEL FOR COVID-19 DEATH USING LDL PARAMETERS

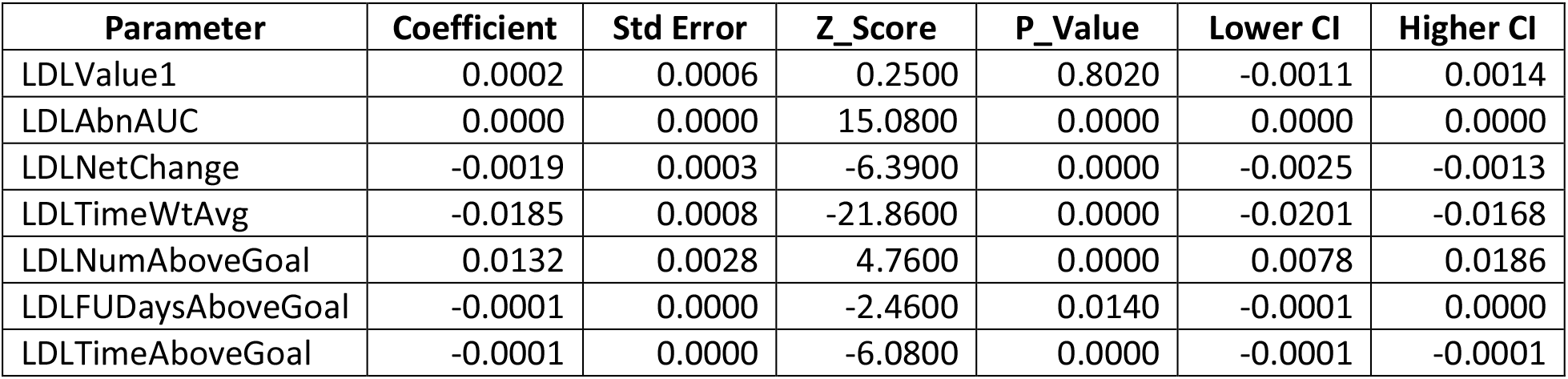
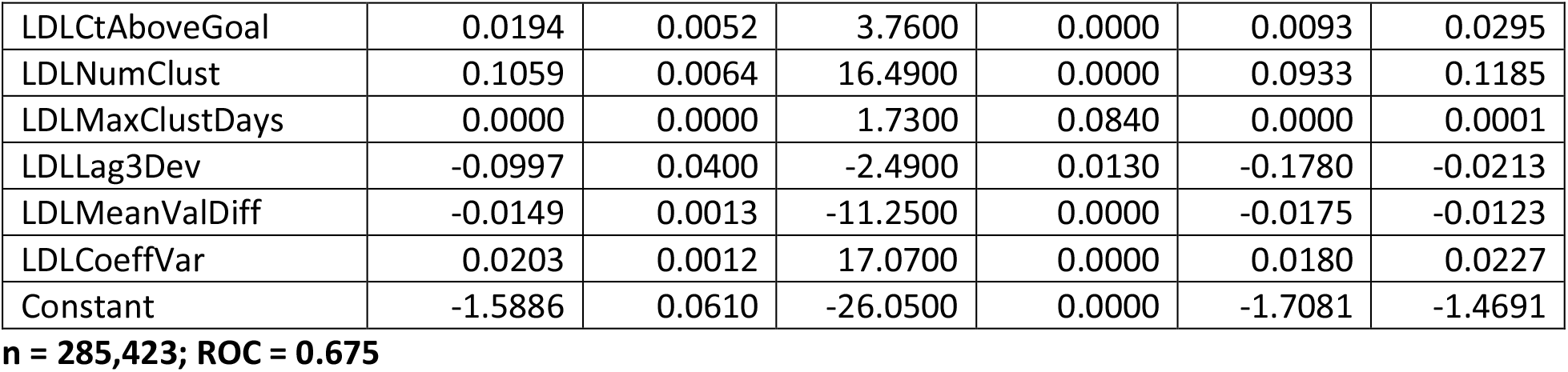

## APPENDIX I: MULTIVARIATE MODEL FOR COVID-19 DEATH USING A1C PARAMETERS

**n = 285,423; ROC = 0.675** APPENDIX I: MULTIVARIATE MODEL FOR COVID-19 DEATH USING A1C PARAMETERS
| Parameter | Coefficient | Std Error | Z_Score | P_Value | Lower CI | Higher CI |
| --- | --- | --- | --- | --- | --- | --- |
| A1cValue1 | -0.0044 | 0.0160 | -0.2800 | 0.7810 | -0.0357 | 0.0269 |
| A1cAbnAUC | 0.0000 | 0.0000 | -5.1200 | 0.0000 | 0.0000 | 0.0000 |
| A1cNetChange | -0.0168 | 0.0071 | -2.3600 | 0.0180 | -0.0307 | -0.0029 |
| A1cTimeWtAvg | 0.0574 | 0.0213 | 2.7000 | 0.0070 | 0.0157 | 0.0991 |
| A1cNumAboveGoal | 0.0016 | 0.0028 | 0.5800 | 0.5600 | -0.0039 | 0.0072 |
| A1cFUDaysAboveGoal | 0.0001 | 0.0000 | 2.4100 | 0.0160 | 0.0000 | 0.0001 |
| A1cTimeAboveGoal | -0.0001 | 0.0000 | -5.0200 | 0.0000 | -0.0001 | -0.0001 |
| A1cCtAboveGoal | 0.0038 | 0.0032 | 1.1600 | 0.2450 | -0.0026 | 0.0101 |
| A1cNumClust | 0.1518 | 0.0083 | 18.2900 | 0.0000 | 0.1355 | 0.1680 |
| A1cMaxClustDays | 0.0002 | 0.0000 | 7.1300 | 0.0000 | 0.0001 | 0.0002 |
| A1cLag3Dev | -0.1333 | 0.1004 | -1.3300 | 0.1840 | -0.3300 | 0.0634 |
| A1cMeanValDiff | -0.2625 | 0.0399 | -6.5800 | 0.0000 | -0.3406 | -0.1843 |
| A1cCoeffVar | 0.0223 | 0.0024 | 9.1000 | 0.0000 | 0.0175 | 0.0270 |
| Constant | -3.5018 | 0.0793 | -44.1600 | 0.0000 | -3.6573 | -3.3464 |

## APPENDIX J: MULTIVARIATE MODEL FOR COVID-19 DEATH USING HDL PARAMETERS

**n = 260,505; ROC = 0.644** APPENDIX J: MULTIVARIATE MODEL FOR COVID-19 DEATH USING HDL PARAMETERS
| Parameter | Coefficient | Std Error | Z_Score | P_Value | Lower CI | Higher CI |
| --- | --- | --- | --- | --- | --- | --- |
| HDLValue1 | -0.0005 | 0.0020 | -0.2600 | 0.7910 | -0.0046 | 0.0035 |
| HDLAbnAUC | 0.0000 | 0.0000 | 1.7800 | 0.0760 | 0.0000 | 0.0000 |
| HDLNetChange | -0.0012 | 0.0010 | -1.2200 | 0.2210 | -0.0032 | 0.0007 |
| HDLTimeWtAvg | 0.0153 | 0.0023 | 6.7000 | 0.0000 | 0.0108 | 0.0198 |
| HDLNumBelowGoal | 0.0189 | 0.0026 | 7.1600 | 0.0000 | 0.0137 | 0.0241 |
| HDLFUDaysBelowGoal | 0.0000 | 0.0000 | -0.8600 | 0.3880 | -0.0001 | 0.0000 |
| HDLTimeBelowGoal | 0.0000 | 0.0000 | -1.5900 | 0.1120 | -0.0001 | 0.0000 |
| HDLCTBelowGoal | 0.0015 | 0.0037 | 0.4200 | 0.6750 | -0.0057 | 0.0088 |
| HDLNumClust | 0.0966 | 0.0062 | 15.5000 | 0.0000 | 0.0844 | 0.1088 |
| HDLMaxClustDays | 0.0001 | 0.0000 | 3.5600 | 0.0000 | 0.0000 | 0.0001 |
| HDLlag3Dev | -0.2636 | 0.0731 | -3.6100 | 0.0000 | -0.4068 | -0.1203 |
| HDLMeanValDiff | -0.0698 | 0.0037 | -18.8000 | 0.0000 | -0.0771 | -0.0625 |
| HDLCoeffVar | 0.0427 | 0.0017 | 24.5700 | 0.0000 | 0.0393 | 0.0461 |
| Constant | -3.9803 | 0.0538 | -73.9700 | 0.0000 | -4.0857 | -3.8748 |

## REFERENCES

1. King JT Jr, Yoon JS, Rentsch CT, et al. Development and validation of a 30-day mortality index based on pre-existing medical administrative data from 13,323 COVID-19 patients: The Veterans Health Administration COVID-19 (VACO) Index. PLoS One. 2020 Nov 11;15(11):e0241825. DOI: 10.1371/journal.pone.0241825.

2. Ioannou GN, Green P, Fan VS, et al. “Development of COVIDVax model to estimate the risk of SARS-CoV-2-related death among 7.6 million US veterans for use in vaccination prioritization,” JAMA Network Open 2021; 4:e214347.

3. Barda N, Riesel D, Akriv A, et a. Developing a COVID-19 mortality risk prediction model when individual-level data are not available. Nature Communications 2020. DOI: 10.1038/s41467-020-18297-9.

4. Hippisley-Cox J, Coupland CAC, Mehta N, et al. Risk prediction of covid-19 related death and hospital admission in adults after covid-19 vaccination: national prospective cohort study. BMJ 2021;374:n2244.

5. Cohn BA, Cirillo PM, Murphy CC, et al. SARS-CoV-2 vaccine protection and deaths among US veterans during 2021. Science 2021. DOI: 10.1126/science.abm0620.

6. Murata GH, Murata AE, Campbell HM, Mcmahon BH, Mao JT. A novel method for handling pre-existing conditions in prediction models for COVID-19 death. Medrxiv 2022. DOI: https://doi.org/10.1101/2022.01.22.22269694

